# Epidemiology of infections with SARS-CoV-2 Omicron BA.2 variant in Hong Kong, January-March 2022

**DOI:** 10.1101/2022.04.07.22273595

**Authors:** Yonatan Mefsin, Dongxuan Chen, Helen S. Bond, Yun Lin, Justin K. Cheung, Jessica Y. Wong, Sheikh Taslim Ali, Eric H. Y. Lau, Peng Wu, Gabriel M. Leung, Benjamin J. Cowling

## Abstract

Hong Kong reported 12,631 confirmed COVID-19 cases and 213 deaths in the first two years of the pandemic but experienced a major wave predominantly of Omicron BA.2.2 in early 2022 with over 1.1 million reported SARS-CoV-2 infections and more than 7900 deaths. Our data indicated a shorter incubation period, serial interval, and generation time of infections with Omicron than other SARS-CoV-2 variants. Omicron BA.2.2 cases without a complete primary vaccination series appeared to face a similar fatality risk to those infected in earlier waves with the ancestral strain.

---

Several variants of concern of SARS-CoV-2 have caused large outbreaks of infection and substantial mortality following the emergence of the ancestral strain in early 2020. The Omicron variant is the most recent variant to have spread globally, since the first detections in Botswana and South Africa in November 2021 [1, 2]. Hong Kong sustained four small epidemic waves in 2020-2021 with 12,631 confirmed cases cumulatively (1.6 cases per 1000 population). Two vaccines became available in early 2021 including the mRNA vaccine BNT162b2 (BioNTech/Fosun Pharma/Pfizer) and the inactivated vaccine CoronaVac (Sinovac), with 70% of the population receiving two doses of vaccination by 31 December 2021. Community outbreaks of Delta and Omicron viruses began around the end of 2021, with Omicron BA.2.2 ultimately dominating a very large fifth wave which peaked in early March 2022. We characterized the epidemiological features of SARS-CoV-2 infections in the fifth wave in Hong Kong.

## COVID-19 epidemics in Hong Kong

Hong Kong has implemented intensive public health and social measures in responses to the COVID-19 pandemic since early 2020 including test-and-trace, isolation of confirmed cases, quarantine of close contacts and inbound travelers in addition to strict travel and border restrictions, and community-wide social distancing measures [3]. These measures had been highly effective in suppressing four previous local outbreaks in 2020 and 2021 but were unable to prevent a large epidemic (wave 5). Since January 2022 a series of stringent social distancing measures have been reimposed in response to the surge of local infections after a lull of reported COVID-19 cases after the end of the fourth wave by May 2021 [4], including suspension of face-to-face classes for all schools from 14 January [5] and school closure for an early summer break from 7 March to 22 April [6], civil servants working from home from 25 January [7], mask mandate in outdoor public places, a ban on group gatherings with more than 2 persons in public places and gatherings at private places involving more than two households, and a suspension of dine-in services after 6pm starting from 10 February [4]. However, the local infections have still reached a record high in wave 5 with over 1.1 million cases officially reported and more than 7600 deaths reported up to date (Figure 1B) while there were in total 12,631 cases and 213 fatalities documented in the first four waves. The daily number of cases detected through RT-PCR and rapid antigen tests peaked on 4 March 2022 after rising exponentially for more than a month with a doubling time of 3.4 days (Figure 1C).

**Figure 1.**
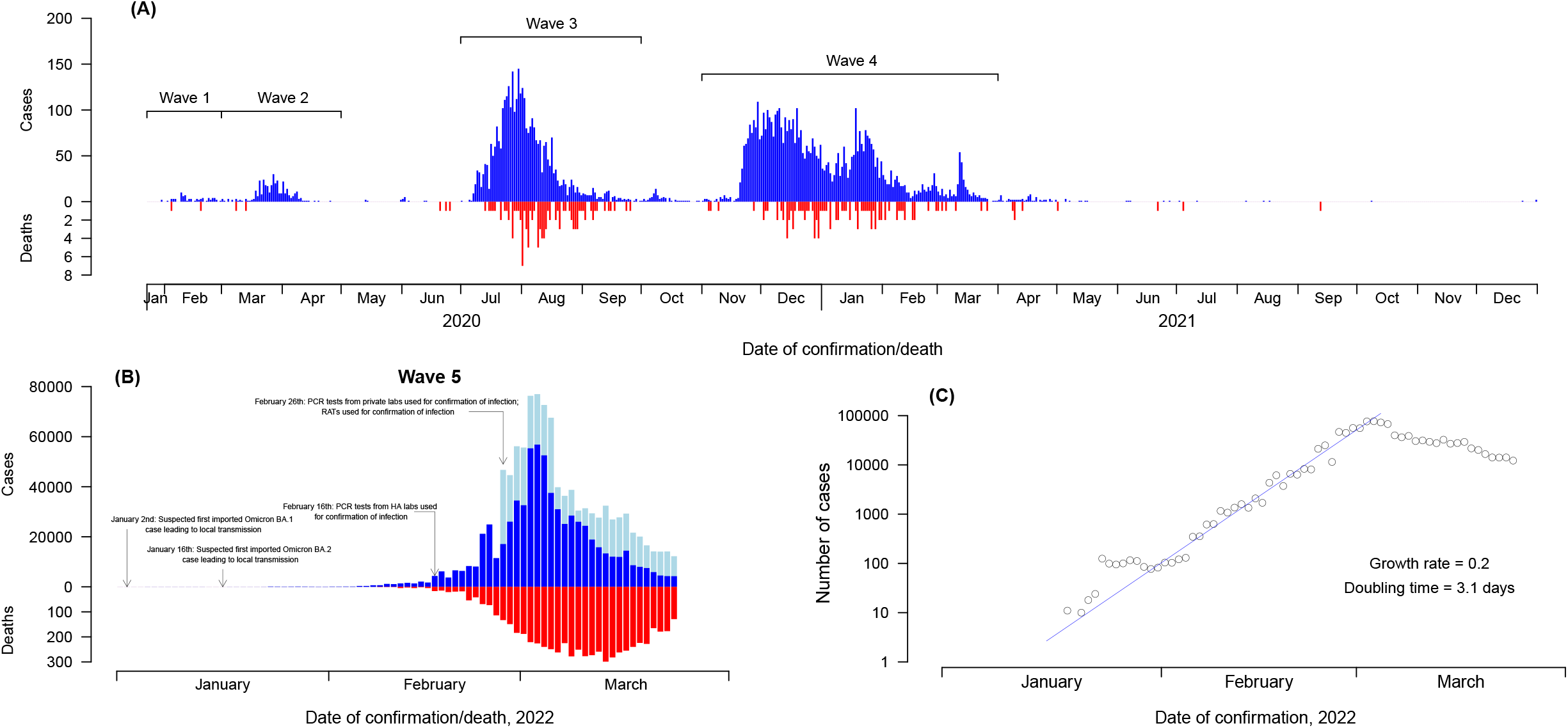
Confirmed COVID-19 local cases and deaths in Hong Kong over the first four epidemic waves (panel A) and wave 5 (panels B and C). Blue and red bars in panels A and B indicated daily numbers of COVID-19 cases confirmed by RT-PCR and daily number of reported deaths, bars in light blue in panel B indicate daily numbers of COVID-19 cases detected by rapid antigen tests (RATs). Panel C indicates the exponential growth of the reported COVID-19 cases in wave 5 before the epidemic peaked. Note: 6 fatal imported cases were also included in panel A, while there were no deaths reported among imported cases in wave 5.

## Epidemiological parameters in wave 5

The Omicron variant of SARS-CoV-2 was first detected in Hong Kong when a within-hotel transmission event of BA.1 was identified in two travelers in hotel quarantine in November 2021 [8]. A small community outbreak of Omicron BA.1 occurred in the community in December 2021, linked initially to restaurants visited by two imported cases in aircrew [9]. The first local Omicron BA.2.2 case was reported in January 2022 linked to a separate transmission event in a quarantine hotel in an arriving traveler that was reaching the end of their 21-day quarantine [10]. Despite stringent public health measures including contact tracing and quarantine not only of close contacts but also close contacts of close contacts, the outbreak of BA.2.2 was not controlled and this strain was responsible for the large epidemic that occurred. Virus sequencing has been done throughout the epidemic, and the last local BA.1 cases and Delta cases were detected in mid-January and early February, respectively, with one sporadic local Delta detection in late March (personal communication, Leo Poon). (personal communication, Leo Poon). The predominance of the Omicron BA.2.2 strain is thought to be a founder effect rather than a particular feature of BA.2.2 as compared to other BA.2 strains (personal communication, Leo Poon).

As of 22 January 2022, 136 locally acquired confirmed Omicron infections were identified in Hong Kong, of which 72% (98/136) had identifiable transmission chains. These cases were linked to four clusters (initiated by imported cases) and the largest cluster comprised 49 cases (Supplementary Figure 1). Of the 98 cases, 34% (33) were Omicron BA.2, 55% (54) were female and 73% were in persons who had completed a primary series of vaccination. All the 98 cases had known exposure information (26% had a single exposure time and the reminder had exposure time intervals) and 80 (82%) reported symptoms at the time of case confirmation (Supplementary Figure 2). Mean (±SD) ages were 44 (±21 for Omicron BA.1 cases and 30 (±21) for Omicron BA.2 cases.

A total of 80 cases (57 BA.1 and 23 BA.2) with known exposure and symptom onset information were included for estimation of the incubation period. The mean (±SD) incubation periods were 4.58 (±1.72) days and 4.42 (±1.42) days for Omicron BA.1 and Omicron BA.2, respectively (Supplementary Table 1). Ninety five percent of patients infected with Omicron BA.1 and BA.2 were estimated to have developed symptoms within 7.7 days and 7.0 days after exposure, respectively (Supplementary Table 1). The gamma distribution had the best fit for BA.1 incubation period distribution.

For serial interval estimation, we used data on 43 symptomatic infector-infectee pairs. The Weibull distribution had the best fit to the serial interval distribution for both Omicron BA.1 and BA.2. For BA.1 (n=30), the estimated mean (±SD) and median serial intervals were 3.30 (±1.95) days and 3.17 days, respectively. For BA.2 (n=13), the estimated mean (±SD) serial interval was 2.72 (±1.51) days, and the median was 2.52 days. All BA.2 estimates were made using gamma distribution accounting for the potential for epidemic phase bias [11] with a growth rate of 0.25, as data were collected during the early growth phase of BA.2 outbreak. We calculated the generation time of BA.1 using data from 45 infector-infectee pairs with known exposure-to-infection time. Using the best-fitted Weibull distribution, the mean and median generation time were 2.36 days (95% confidence interval (CI): 2.01, 2.77) and 2.38 days (95% CI: 2.01, 2.80) for BA.1, respectively.

## Case-fatality-risk across epidemic waves

Among the 6320 deaths in cases confirmed in wave 5 by 23 March 2022, 103 were reported on or before 15 February (early period of wave 5) when all COVID-19 cases were only confirmed through RT-PCR conducted by the Public Health Laboratory Services of Hong Kong Government, same as in earlier waves (Figure 1, Supplementary material). We classified cases as having a primary series of vaccination if they had received two doses for mRNA BNT162b2 (BioNTech/Fosun Pharma) and/or inactivated CoronaVac (Sinovac) with the most recent dose at least 14 days earlier. After accounting for unresolved outcomes in some individuals, we estimated that the age-specific case-fatality-risk (CFR) for cases without completion of a primary series of vaccination in wave 5 was comparable to those confirmed in waves 1-4 across all age groups (Figure 2, Supplementary Table 2), while the highest fatality risk was observed in patients aged 80 years and above (24.9%, 95% CI: 20.9, 29.3) in waves 1-4, similar to the unvaccinated at the same age (21.7%, 95% CI: 17.1, 26.8) from wave 5 (Figure 2). The CFR for cases in persons ≥80 years who had not completed a primary series of vaccination in wave 5 appeared to be approximately double the risk in persons in the same age group who had completed a primary series (11.1%, 95% CI: 4.2, 22.6), and the pattern remained the same for cases at 64-79 years of age (Figure 2, Supplementary Table 2). The 7 deaths that occurred in older adults aged ≥65 years with a complete primary series of vaccination were all from persons who had received two doses of CoronaVac.

**Figure 2.**
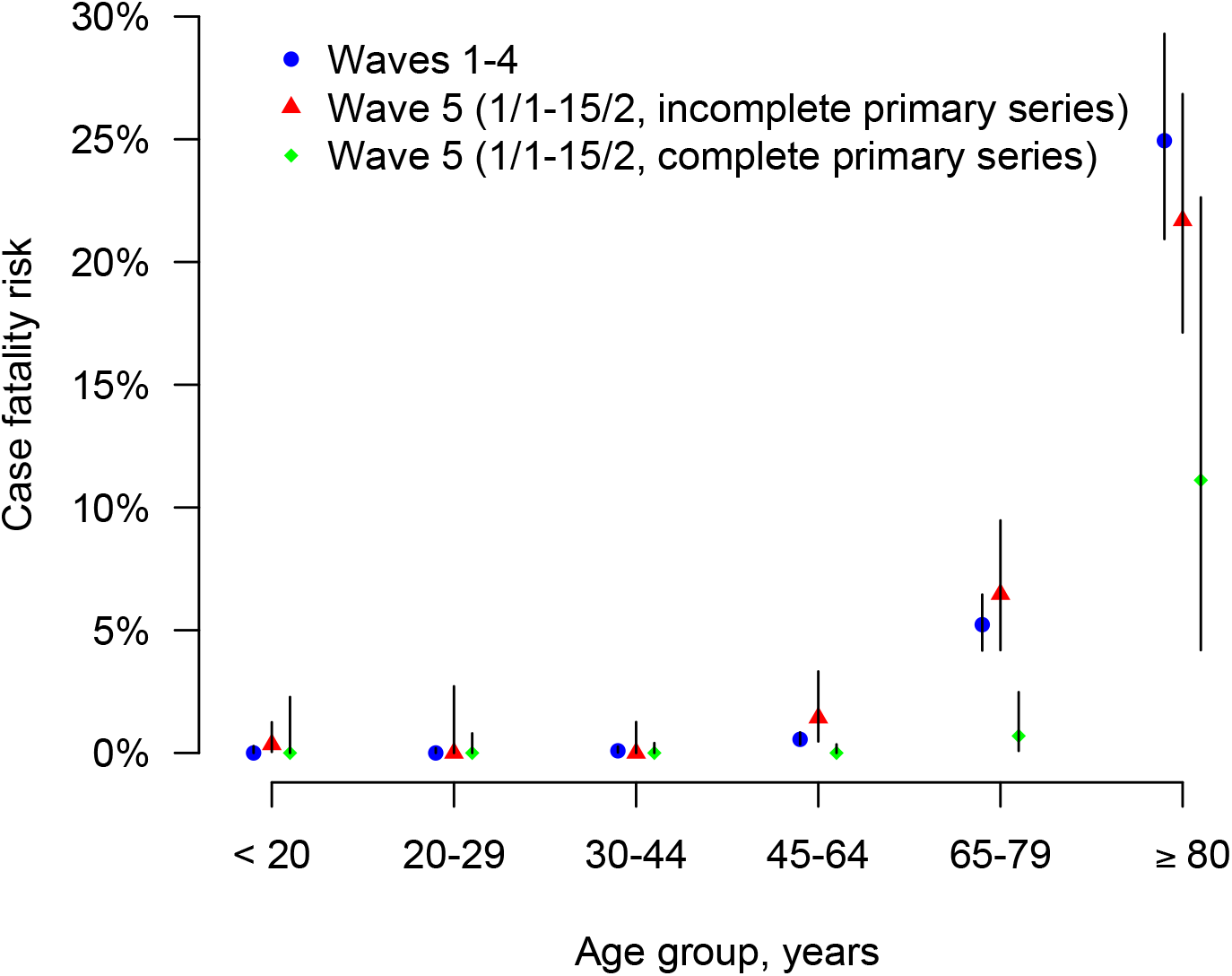
Age-stratified estimates of the case-fatality-risk of COVID-19 in epidemic waves 1-4 and wave 5 in Hong Kong by vaccination status. Note: cases were classified as having a complete primary series if receiving 2 or more doses of COVID-19 vaccines at least two weeks before symptom onset (for symptomatic cases) or at least three weeks before laboratory confirmation of the infection (for symptomatic cases with a missing onset date or asymptomatic cases), otherwise as having an incomplete primary series.

## Discussion

The SARS-CoV-2 Omicron BA.2 variant has caused more than 1.1 million confirmed cases in Hong Kong (mid-year population 7.4 million) within three months in early 2022, 100-fold higher than all the cases confirmed over four epidemic waves in the previous two years. Not every infection would become a laboratory-confirmed case for various reasons [12]. The peak in the epidemic in early March despite no major change in social distancing measures is likely indicative of a sufficient number of infections to create herd immunity, at least temporarily, with subsequent infections “overshooting” that threshold [13]. Despite stringent public health and social measures, Omicron BA.2.2 spread rapidly with a doubling time of 3.4 days (Figure 1C), quickly outpacing the capacity in test-and-trace, isolation, and quarantine, leading to a large number of patients in need of admission with limited availability of resources. The relatively shorter serial interval and generation time of the Omicron BA.2 subvariant in Hong Kong (Supplementary Figure 3, Supplementary Table 1) compared to earlier variants [14] would have been one of the factors contributing to faster spread in the population along with the higher intrinsic transmissibility (e.g. higher reproductive number).

The high number of deaths in Hong Kong’s fifth wave can be attributed to the high incidence of infections within a short period of time, and the low level of vaccination coverage in older adults. While the overall vaccine coverage at the start of the fifth wave was 70%, in persons ≥65 years and ≥80 years only 50% and 20% had completed a primary series of vaccination. Vaccine hesitancy in older adults occurred for a variety of reasons [15]. Among all the deaths with age recorded up to 23 March, 92.5% (5844/6318) occurred in persons ≥65 years of age and 70.8% (4472/6318) in persons ≥80 years. Infections with the Omicron variant demonstrated attenuated pathogenesis in animal models [16] and milder severity in South Africa [17, 18] and elsewhere [19–21]. However, we found a similar fatality risk for unvaccinated cases in the early part of our fifth wave compared to earlier waves, indicating that the intrinsic severity of BA.2 may not be much lower than the ancestral strain if at all. We found that cases in persons who had a complete primary series at age of 65-79 years or ≥80 years would have a much lower fatality risk (Figure 2, Supplementary Table 2). In a separate study, we estimated very high vaccine effectiveness against severe disease in older adults ≥60 years of age who received either three doses of the inactivated vaccine CoronaVac or two doses of BNT162b2 [22]. For adults ≥60 years of age, the World Health Organization recommend that three doses of inactivated vaccine are needed [23]. Our estimates of the CFR might slightly overestimate the fatality risk of Omicron in Hong Kong given there was a lack of information on the type of virus variant for all individual cases and a small number of cases including deaths from clusters of Delta infections might have been included in the analysis.

## Conclusions

A higher transmission potential was indicated for Omicron variants particularly the BA.2 subvariant in Hong Kong. Cases with a complete primary vaccination series had a much lower fatality risk compared with those without vaccination or with an incomplete primary series of vaccination. Our findings highlighted the importance of achieving a high coverage of vaccination especially in older adults and the need to reassess public health and social measures in control of epidemics in response to a more transmissible SARS-CoV-2 variant in the future.

## Data Availability

All data produced in the present study are available upon reasonable request to the authors.

## ACKNOWLEDGEMENTS

The authors thank Julie Au, Chloe Chui, Caitriona Murphy, Faith Ho, and Dillon Adam for technical support.

## AUTHOR CONTRIBUTIONS

YM, PW and BJC conceived the study. YM, DC, HB, YL, JKC, JYW, STA and EHY collected the data and conducted the analysis. YM and PW drafted the manuscript. All authors critically reviewed and revised the manuscript and approved the final version.

## FUNDING

This project was supported by the Collaborative Research Scheme (project no. C7123-20G) of the Research Grants Council of the Hong Kong SAR Government, the commissioned Health and Medical Research Fund from the Food and Health Bureau of the Hong Kong SAR Government (grant no. CID-HKU2), and AIR@InnoHK administered by Innovation and Technology Commission. BJC is supported by a RGC Senior Research Fellow Scheme grant (HKU SRFS2021-7S03) from the Research Grants Council of the Hong Kong Special Administrative Region, China.

## CONFLICTS OF INTEREST

BJC consults for AstraZeneca, Fosun Pharma, GSK, Moderna, Pfizer, Roche and Sanofi Pasteur. The authors report no other potential conflicts of interest.

## Notes

### Author Declarations

Ethics approval for this study was obtained from the Institutional Review Board of The University of Hong Kong/Hospital Authority Hong Kong West Cluster (HKU/HA HKW IRB). Data collection and analysis were part of a continuing public health outbreak investigation. Informed consent was not required.

