## Supplementary File for "Epidemiology of infections with SARS-CoV-2 Omicron BA.2 variant in Hong Kong, January-March 2022"

**Supplementary materials**

**Data sources**

We obtained all confirmed cases with SARS-CoV-2 infections in Hong Kong from the Department of Health and the Hospital Authority of the Government of Hong Kong Special Administrative Region from 23 January 2020 to 17 March 2022. Case information includes age, sex, time of symptom onset, sample collection, laboratory confirmation, report, admission, discharge and death, severity status (mild-to-moderate, severe or critical, fatal), SpO2 levels, time and vaccination and type of vaccine for each dose. We also collected contact tracing data published by the Government or from media reports that quoted government statements on some of the confirmed case clusters [1]. The contact tracing data provided information on demographics (age and sex), date of exposure, symptom onset date, case category (symptomatic /asymptomatic), type of SARS-CoV-2 variant, and vaccination status. The confirmed cases in a cluster either had a single exposure date or a reported interval of exposure defined by the earliest and latest dates of exposure to an index case. In Hong Kong a COVID-19 case can be confirmed by the real-time reverse transcription-polymerase chain reaction (real-time RT-PCR) assay (since the emergence of SARS-CoV-2 in 2020) or by a rapid antigen test (RAT) since 26 February 2022.

**Laboratory confirmation of COVID-19 in Hong Kong**

|  | Waves 1-4 | Wave 5 (1 Jan 2022 till now) | | |
| --- | --- | --- | --- | --- |
|  | (1 Jan 2020-31 Dec 2021) | 1 Jan-15 Feb | 16 Feb-25 Feb | 26 Feb and after |
| RT-PCR, PHLS |  |  |  |  |
| RT-PCR, HA labs |  |  |  |  |
| RT-PCR, commercial labs |  |  |  |  |
| Rapid antigen test (RAT) |  |  |  |  |

Note: PHLS: The Public Health Laboratory Services Branch of Centre for Health Protection provides clinical diagnostic and public health laboratory services to the public and private health sectors for both patient care and public health functions. HA: The Hospital Authority is a statutory body managing all the government hospitals and institutes in Hong Kong.

**Epidemiological parameters definition**

We define that the incubation period is the time interval between initial contact with a confirmed SARS-CoV-2 case and symptom onset, serial interval is the time interval between symptom onset of the infector and the infectee in a transmission pair, and generation time is the time interval between exposure time of the infector and the infectee in a transmission pair.

**Transmission pair construction**

We constructed infector-infectee transmission pairs with laboratory-confirmed SARS-CoV-2 omicron infection using contact tracing data published on the website of the Department of Health, the Government of the Hong Kong Special Administrative Region, and media reports that quoted government statements.

We constructed transmission pairs for estimating mean serial interval and generation time if the infector had direct contact with its infectee. A transmission pair was defined as two confirmed COVID-19 cases identified in the epidemiologic investigation by showing a clear epidemiologic link with each other, which is the infector needs to have direct contact with the infectee. The ‘infector’ was defined as the primary case, with an identified source of exposure occurring prior to their encounter with the ‘infectee’. The ‘infectee’ was defined as the secondary case whose exposure was solely by the infector. In the same chain of transmission, the same infector could generate more than one pair if they infected more than one individual. In a large case cluster with multiple generations of transmission, an infectee in a transmission pair might also be an infector in another pair.

For serial interval calculation, as there is established evidence of asymptomatic/ presymptomatic transmission of SARS-CoV-2 [2, 3], infector-infectee pairs were determined regardless of the order of their onset dates. The infectors-infectees order were determined by who were first exposed to suspected source of infection and induced further infection spread to his/her own network, especially in the case of household or workplace transmission setting. However, to be conservative, in a cluster where the serial interval between index case to off-spring case was larger than 6 days [4], such transmission pairs were excluded as this was suggestive of possible intermediate transmission.

For exposure time that was not explicitly recorded in the government document, we constructed exposure interval based on the following assumptions:

1.	The latest exposure time (lower bounds of the exposure interval) is symptom onset date or isolation/quarantine date or confirmation date (if asymptomatic) of the infector

2.	The earliest exposure time (upper bounds of the exposure interval) is the infector’s exposure date if the infector was a local family member or the infector’s arrival date if the infector was an imported family member.

**Statistical analysis**

We fitted the parametric distributions of Lognormal, Weibull, and Gamma models to the time intervals data and estimated the distributions of incubation period, serial interval and generation time using the maximum likelihood method [5-7]. We accounted for the interval censoring of exposure windows in estimation of the incubation period and generation time. The best fitted model was determined by the smallest value of Akaike’s Information Criterion. Estimates of the mean, standard deviation (SD), median and 95% percentiles were derived from the models. The corresponding 95% confidence intervals (CIs) of each estimate were constructed using the parametric bootstrap method with 1000 bootstrapped samples. Because data on serial interval included zero value, we shifted data by adding 1 days to each serial interval so that we could fit distributions. Furthermore, Omicron BA.2 parameters estimations were also made using gamma distribution accounting for the sampling bias that could happen when parameters estimation made using data collected during the initial growth phase of the outbreak [8]. In our calculation, sampling bias correction was made based on the computed exponential growth rate (r=0.25) of BA.2 using data collected from 15-23 January 2022.

**Figures and Tables**

**Supplementary Figure 1.** Transmission chains by transmission settings of SARS-CoV-2 Omicron infections associated with four imported cases in Hong Kong, 31 December 2021 to 22 January (n=98).

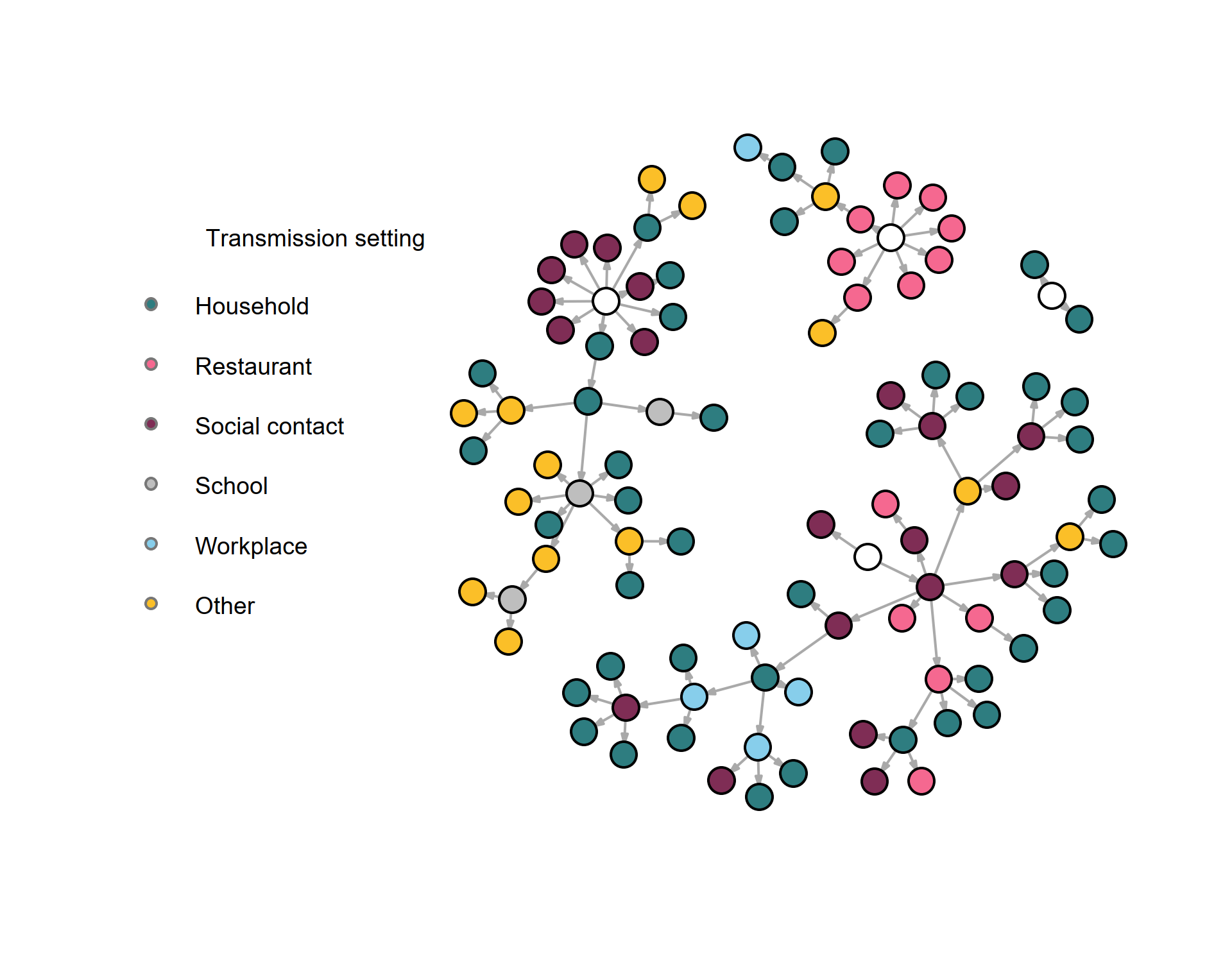

**Supplementary Figure 2**: Cases (n=98) studied to estimate incubation period, serial interval and generation time of SARS-CoV-2 Omicron variant. Figure A refers to symptomatic cases and Figure B refers to asymptomatic cases. In each row, red shaded point indicates the dates of onset date, orange shaded point indicates confirmation rate, blue shaded area indicates the period of exposure and that in grey indicates the incubation period.

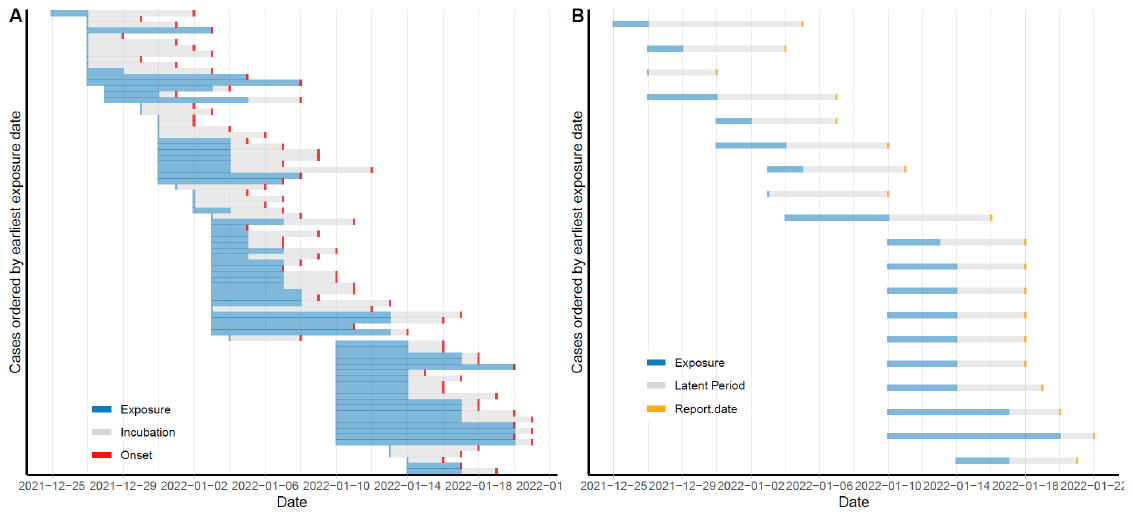

**Supplementary Figure 3.** The estimated distributions of the incubation period, serial interval and generation time for infections occurred in infections with Omicron variant BA.1 and Omicron BA.2 in wave 5 in Hong Kong.

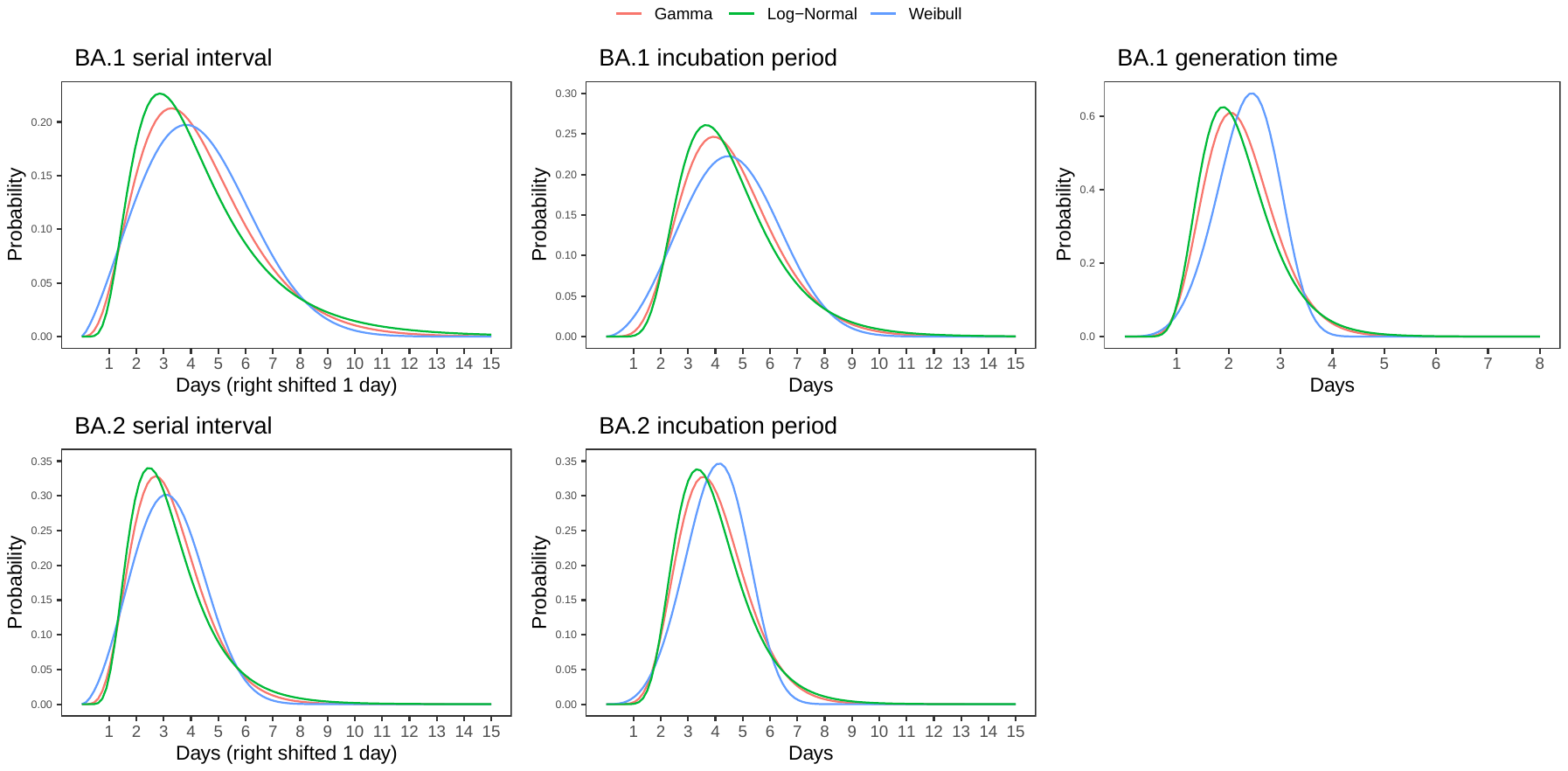

**Supplementary Table 1**. **Estimated mean and percentiles of incubation period, serial interval and generation time of infections with SARS-CoV-2 Omicron BA.1 and BA.2 subvariants, 31 December 2021 to 22 January 2022, Hong Kong.**

| **Omicron subvariants** | **Parameters (sample size)** | **Best fitted distribution** | **Mean (95% CI)** | **SD (95% CI)** | **Median (95%CI)** | **95 percentile (95% CI)** |
| --- | --- | --- | --- | --- | --- | --- |
| BA.1 | Incubation period (57) | Gamma | 4.58  (4.1 - 5.08) | 1.72  (1.32 - 2.07) | 4.38  (3.88 - 4.87) | 7.73  (6.72 - 8.71) |
|  | Serial Interval (30) | Weibull | 3.30  (2.65-4.01) | 1.96  (1.42-2.41) | 3.15  (2.49-3.92) | 6.76  (5.36-8.16) |
|  | Generation Time (45) | Weibull | 2.36  (2.01 – 2.77) | 0.59  (0.38 – 0.90) | 2.38  (2.01-2.80) | 3.28  (2.79-3.97) |
| BA.2 | Incubation period (23) | Weibull | 4.03  (3.19, 4.80) | 1.12 (0.46, 1.51) | 4.05  (3.21, 4.84) | 5.82  (4.37, 6.76) |
|  |  | Gamma* | 4.42  (3.40-5.21) | 1.42  (0.36-1.99) | 4.27  (3.29-5.02) | 6.93  (4.85-8.80) |
|  | Serial Interval (13) | Weibull | 2.23  (1.53-2.90) | 1.26  (0.72-1.63) | 2.17  (1.46-2.91) | 4.40  (2.99-5.31) |
|  |  | Gamma* | 2.72  (1.80-3.88) | 1.51  (0.76-2.43) | 2.52  (1.68-3.55) | 5.50  (3.31-8.25) |

* Estimation accounted for the potential epidemic phase bias since Omicron BA.2 were data collected during the early growth phase of BA.2 outbreak. The exponential growth rate was estimated to be 0.25 based on data collected from 15-23 January 2022.

**Supplementary Table 2. Estimated case-fatality-risk among the COVID-19 cases confirmed in the first 1-4 waves in comparison to cases with and without complete primary series who were identified in the early period of wave 5 (1 January – 15 February 2022) in Hong Kong.**

|  | **Case-fatality-risk, mean (95% confidence interval)** | | | | | |
| --- | --- | --- | --- | --- | --- | --- |
|  | **< 20 years** | **20-29 years** | **30-44 years** | **45-64 years** | **65-79 years** | **80+ years** |
| **Wave 1 to 4** | 0  (0, 0.28) | 0  (0, 0.19) | 0.09  (0.02, 0.26) | 0.55  (0.35, 0.84) | 5.23  (4.17, 6.45) | 24.94  (20.93, 29.30) |
| **Wave 5**  **(Complete primary series) ^‡^** | 0  (0, 2.28) | 0  (0, 0.80) | 0  (0, 0.40) | 0  (0, 0.35) | 0.69  (0.08, 2.48) | 11.11  (4.19, 22.63) |
| **Wave 5**  **(Incomplete primary series) ^‡^** | 0.35  (0.04, 1.25) | 0  (0, 2.72) | 0  (0, 1.26) | 1.44  (0.47, 3.32) | 6.47  (4.19, 9.47) | 21.69  (17.13, 26.84) |

^‡^ Cases with complete primary series are individuals who have received at least 2 doses of COVID-19 vaccines before confirmation of infection, and cases with incomplete vaccinated refer to those without receiving any vaccine or with only one dose of vaccine.
